# Analysis of 200,000 exome-sequenced UK Biobank subjects fails to identify genes influencing probability of developing a mood disorder resulting in psychiatric referral

**DOI:** 10.1101/2021.01.07.20249042

**Authors:** David Curtis

**Affiliations:** UCL Genetics Institute, UCL, Darwin Building, Gower Street, London WC1E 6BT; Centre for Psychiatry, Queen Mary University of London, Charterhouse Square, London EC1M 6BQ

**Author notes:** Correspondence: David Curtis.

**Keywords:** Depression, exome, UK Biobank, gene

## Abstract

**Background:** Depression is moderately heritable but there is no common genetic variant which has a major effect on susceptibility. A previous analysis of 50,000 subjects failed to implicate any genes or sets of genes associated with risk of affective disorder requiring specialist treatment. A large exome-sequenced dataset is now available.

**Methods:** Data from 200,632 exome-sequenced UK Biobank participants was analysed. Subjects were treated as cases if they had reported having seen a psychiatrist for “nerves, anxiety, tension or depression”. Gene-wise weighted burden analysis was performed to see if there were any genes or sets of genes for which there was an excess of rare, functional variants in cases.

**Results:** There were 22,886 cases and 176,486 controls. There were 22,642 informative genes but no gene or gene set produced a statistically significant result after correction for multiple testing. None of the genes or gene sets with the lowest p values appeared to be a biologically plausible candidate.

**Limitations:** The phenotype is based on self-report and the cases are likely to somewhat heterogeneous. Likewise, it is expected that some of the subjects classed as controls will in fact have suffered from depression or some other psychiatric diagnosis.

**Conclusions:** The results conform exactly with the expectation under the null hypothesis. It seems unlikely that the use of common, poorly defined phenotypes will produce useful advances in understanding genetic contributions to affective disorder and it might be preferable to focus instead on obtaining large exome-sequenced samples of conditions such as bipolar 1 disorder and severe, recurrent depression.

## Introduction

Genetic investigations of diseases such as schizophrenia and Alzheimer’s disease have resulted in the identification of individual genes affecting susceptibility and such results provide helpful information which may eventually result in advances in therapy (Curtis and Bandyopadhyay, 2021; Mukai et al., 2019). By contrast, genome wide association studies of less severe mental illnesses such as depression have failed to reveal genes with a substantial effect on risk, while a study of 50,000 exome-sequenced UK Biobank subjects was completely unsuccessful in highlighting any genes influencing the probability of being referred to a specialist for treatment of a mood disorder (Curtis, 2021; Howard et al., 2019). The latter study concluded that it was unlikely that specific genes would be implicated until far larger samples become available.

As explained previously, the UK Biobank is a resource of 500,000 British volunteers for whom variable amounts of phenotypic information is available (http://www.ukbiobank.ac.uk/about-biobank-uk/) (Curtis, 2021). One of the measures completed by almost all subjects was an item at the initial assessment, consisting of a touchscreen question asking, “Have you ever seen a psychiatrist for nerves, anxiety, tension or depression?” to which 497,000 answered either Yes or No. This item was chosen as the measure of interest because most subjects had responded and because requiring referral to a specialist might reasonably be taken as a measure of having some significant degree of psychiatric morbidity. Weighted burden analysis of rare variants within genes was carried out on the 50,000 subjects for whom exome sequence data was available, with variants which were more likely to have functional effects and/or which were rarer receiving higher weights. In this sample there was no gene for which there was a statistically significant association between the weighted burden score and probability of referral for specialist psychiatric treatment. UK Biobank has now increased the number of subjects for whom exome sequence is available to 200,000 and we report here the results of applying the same analytic approach to this expanded dataset.

## Methods

The same approach was followed as previously fully described (Curtis, 2021). The UK Biobank dataset was downloaded along with the variant call files for 200,632 subjects who had undergone exome-sequencing and genotyping by the UK Biobank Exome Sequencing Consortium using the GRCh38 assembly with coverage 20X at 95.6% of sites on average (Szustakowski et al., 2020). UK Biobank had obtained ethics approval from the North West Multi-centre Research Ethics Committee which covers the UK (approval number: 11/NW/0382) and had obtained informed consent from all participants. The UK Biobank approved an application for use of the data (ID 51119) and ethics approval for the analyses was obtained from the UCL Research Ethics Committee (11527/001). All variants were annotated using the standard software packages VEP, PolyPhen and SIFT (Adzhubei et al., 2013; Kumar et al., 2009; McLaren et al., 2016). To obtain population principal components reflecting ancestry, version 2.0 of *plink* (https://www.cog-genomics.org/plink/2.0/) was run with the options *--maf 0.1 --pca 20 approx* (Chang et al., 2015; Galinsky et al., 2016).

The phenotype was determined according to how participants had responded in their initial assessment to the touchscreen question: “Have you ever seen a psychiatrist for nerves, anxiety, tension or depression?” Those answering “Yes” were taken to be cases and those answering “No” controls. No attempt was made to screen out controls who might have had some other psychiatric diagnosis.

SCOREASSOC was used to carry out a weighted burden analysis to test whether, in each gene, sequence variants which were rarer and/or predicted to have more severe functional effects occurred more commonly in cases than controls. Attention was restricted to rare variants with minor allele frequency (MAF) <= 0.01 in both cases and controls. As previously described, variants were weighted by MAF so that variants with MAF=0.01 were given a weight of 1 while very rare variants with MAF close to zero were given a weight of 10. Variants were also weighted according to their functional annotation, with weights ranging from 1 for intergenic variants to 40 for variants predicted to cause complete loss of function in the gene. Additionally, 10 was added to the weight if the PolyPhen annotation was possibly or probably damaging and also if the SIFT annotation was deleterious. The full range of annotations and their weights is shown in Table 1. The weight due to MAF and the weight due to functional annotation were then multiplied together to provide an overall weight for each variant. For each subject a gene-wise weighted burden score was derived as the sum of the variant-wise weights, each multiplied by the number of alleles of the variant which the given subject possessed. For variants on the X chromosome, hemizygous males were treated as homozygotes. Variants were excluded if there were more than 10% of genotypes missing in the cases or controls or if the heterozygote count was smaller than both homozygote counts in the cases or controls. If a subject was not genotyped for a variant then they were assigned the subject-wise average score for that variant.

**Table 1.** The table shows the weight which was assigned to each type of variant as annotated by VEP, Polyphen and SIFT (Adzhubei et al., 2013; Kumar et al., 2009; McLaren et al., 2016).

| **VEP / SIFT / Polyphen annotation** | **Weight** |
| --- | --- |
| intergenic_variant | 1 |
| feature_truncation | 3 |
| regulatory_region_variant | 3 |
| feature_elongation | 3 |
| regulatory_region_amplification | 3 |
| regulatory_region_ablation | 3 |
| TF_binding_site_variant | 3 |
| TFBS_amplification | 3 |
| TFBS_ablation | 3 |
| downstream_gene_variant | 3 |
| upstream_gene_variant | 3 |
| non_coding_transcript_variant | 3 |
| NMD_transcript_variant | 3 |
| intron_variant | 3 |
| non_coding_transcript_exon_variant | 3 |
| 3_prime_UTR_variant | 10 |
| 5_prime_UTR_variant | 5 |
| mature_miRNA_variant | 5 |
| coding_sequence_variant | 5 |
| synonymous_variant | 5 |
| stop_retained_variant | 5 |
| incomplete_terminal_codon_variant | 5 |
| splice_region_variant | 5 |
| protein_altering_variant | 10 |
| missense_variant | 10 |
| inframe_deletion | 15 |
| inframe_insertion | 15 |
| transcript_amplification | 15 |
| start_lost | 30 |
| stop_lost | 30 |
| frameshift_variant | 40 |
| stop_gained | 40 |
| splice_donor_variant | 40 |
| splice_acceptor_variant | 40 |
| transcript_ablation | 20 |
| SIFT deleterious | 10 |
| PolyPhen possibly damaging | 10 |
| PolyPhen probably damaging | 10 |

For each gene, a ridge regression analysis was carried out with lamda=1 to test whether the gene-wise variant burden score was associated with the psychiatric phenotype, including the first 20 population principal components and sex as covariates. The statistical significance is summarised as a signed log p value (SLP) which is the log base 10 of the associated p value, given a positive sign if the score is higher in cases and negative if it is higher in controls.

Gene set analyses were carried out using the 1454 “all GO gene sets, gene symbols” pathways as listed in the file *c5.all.v5.0.symbols.gmt* downloaded from the Molecular Signatures Database at http://www.broadinstitute.org/gsea/msigdb/collections.jsp (Subramanian et al., 2005). For each set of genes, the natural logs of the gene-wise p values were summed according to Fisher’s method to produce a chi-squared statistic with degrees of freedom equal to twice the number of genes in the set. The p value associated with this chi-squared statistic was expressed as a minus log10 p (MLP) as a test of association of the set with the mental illness phenotype.

## Results

There were 22,886 cases who answered positively to the question about having seen a psychiatrist for “nerves, anxiety, tension or depression” and 176,486 controls. There were 22,642 genes for which there were qualifying variants and the QQ plot for the SLPs obtained for each gene is shown in Figure 1. This shows that the test is well-behaved and that the SLPs conform exactly to what one would expect under the null hypothesis that there are no genes for which an increased burden of rare, functional variants affects the risk of mental illness.

**Figure 1.**
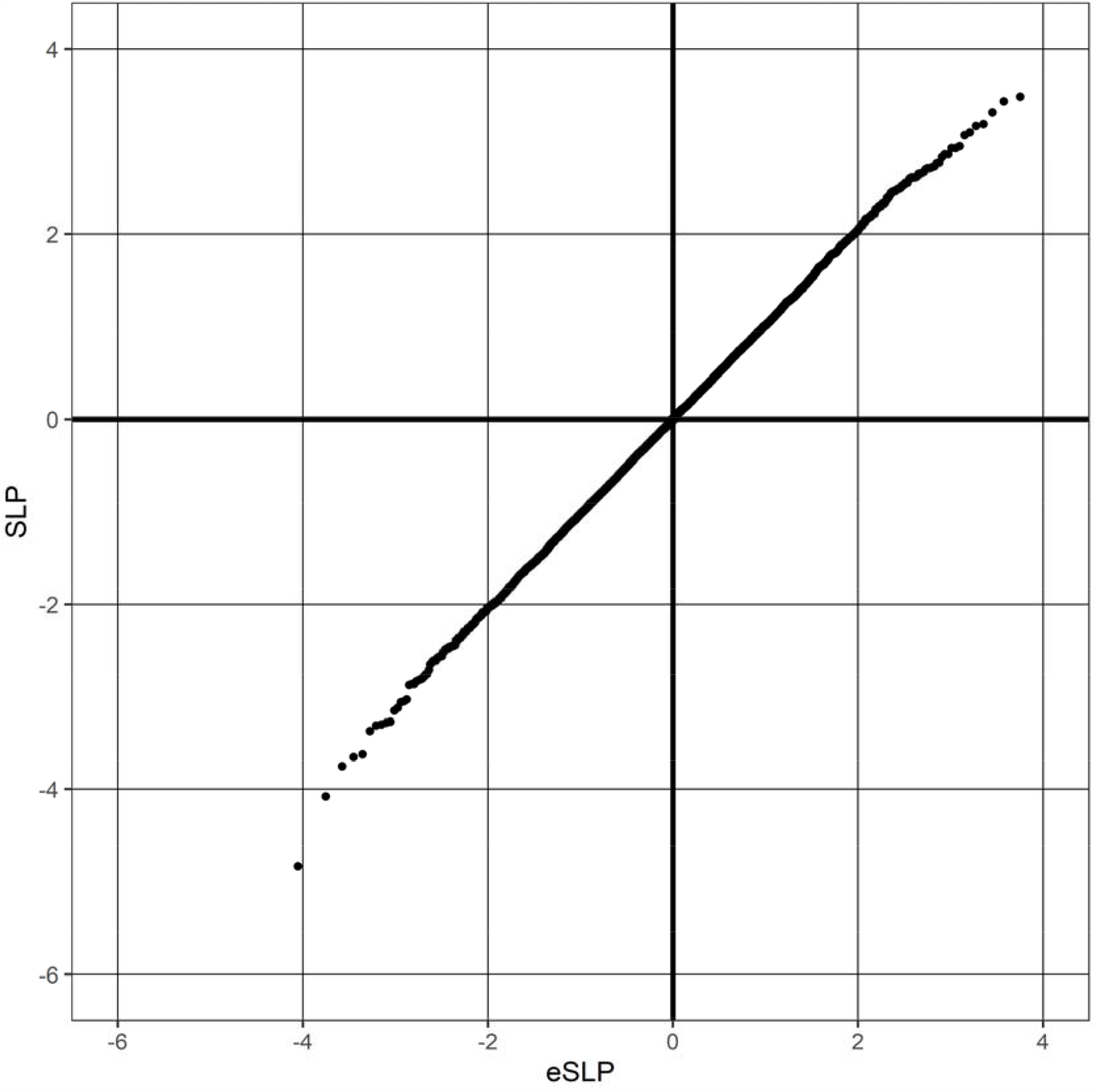
QQ plot of SLPs obtained for weighted burden analysis of 22,642 genes for association with referral for psychiatric treatment showing observed against expected SLP for each gene.

**Table 2.** Genes with absolute value of SLP exceeding 3 or more (equivalent to p<0.001) for test of association of weighted burden score with referral for psychiatric treatment.

| Gene symbol | SLP | Gene name |
| --- | --- | --- |
| *CLDND1* | 3.95 | Claudin Domain Containing 1 |
| *DUSP16* | 3.48 | Dual Specificity Phosphatase 16 |
| *LOC101928847* | 3.43 | Uncharacterized LOC101928847 |
| *EFTUD2* | 3.31 | Elongation Factor Tu GTP Binding Domain Containing 2 |
| *LOC105377103* | 3.19 | Uncharacterized LOC105377103 |
| *EED* | 3.17 | Embryonic Ectoderm Development |
| *HIST2H3D* | 3.07 | H3 Clustered Histone 13 |
| *JADE3* | -3.05 | Jade Family PHD Finger 3 |
| *APBA2* | -3.06 | Amyloid Beta Precursor Protein Binding Family A Member 2 |
| *LOC105370163* | -3.27 | Uncharacterized LOC105370163 |
| *CCDC160* | -3.28 | Coiled-Coil Domain Containing 160 |
| *ADHFE1* | -3.31 | Alcohol Dehydrogenase Iron Containing 1 |
| *MIR6720* | -3.37 | MicroRNA 6720 |
| *IFRD2* | -3.62 | Interferon Related Developmental Regulator 2 |
| *MAGEA8-AS1* | -3.65 | MAGEA8 Antisense RNA 1 |
| *LOC112267907* | -3.75 | Uncharacterized LOC112267907 |
| *COX18* | -4.08 | Cytochrome C Oxidase Assembly Factor COX18 |
| *MIR4695* | -4.83 | MicroRNA 4695 |

Table 1 shows the results for all genes with an absolute value of SLP >= 3 (equivalent to p<=0.001). By chance, from 22,642 genes one would expect 11 to have SLP greater than 3 and 11 to have SLP less than −3, whereas the actual numbers are 7 and 13. Applying a Bonferroni correction to test for genome-wide statistical significance would yield a threshold of log10(22,028/0.05) = 5.6 for the absolute value of the SLP and no gene achieves this. The genes listed in the table are involved in a wide variety of different functions but none appears to be a plausible biological candidate for affecting susceptibility to depression or anxiety.

There is no gene set which would be statistically significant after a Bonferroni correction for the number of sets tested. The most significant set was TRANSFORMING GROWTH FACTOR BETA RECEPTOR SIGNALING PATHWAY (MLP = 4.07) which contains 36 genes. Neither this set nor the others with relatively high MLPs appear to be biologically plausible candidates.

The SLPs for all genes and the MLPs for all gene sets are provided in supplementary tables S1 and S2. Discussion

## Discussion

Even with the considerably larger sample, the results obtained from this study are as completely negative as were the results from the previous analysis of 50,000 exomes. None of the results is formally significant and even the genes which are ranked highest and lowest do not include any which could be regarded as being biologically plausible candidates. This contrasts with results obtained when applying similar methods to considerably smaller sample sizes of other complex phenotypes, comprising schizophrenia, late onset Alzheimer’s disease, BMI and hyperlipidaemia (Curtis, 2020b, 2020a; Curtis et al., 2019, 2018). For these phenotypes it was possible to identify gene sets which were statistically significant and a small number of biologically plausible genes which were either genome-wide significant or which at least had uncorrected p values less than 0.001. In the present dataset there is no suggestion of such a signal and the results follow a completely random distribution.

As discussed previously, the fact that the results are negative may arise from a number of factors. Obviously, the answer to a single touch screen question results in a poorly defined phenotype. Nevertheless, referral to a specialist might be taken as an indicator of significant psychiatric morbidity, more so than a simple self-report of anxiety or depression and/or being prescribed medication. One might expect that the bulk of the cases might have a mix of anxiety and depression related diagnoses and studies using common genetic variants suggest a considerable overlap for genetic contributions to these conditions (Purves et al., 2019). However the overall heritability of depression is lower than for schizophrenia and it is possible that rare genetic variants with major effect sizes make a smaller overall contribution to liability. It may be that some variants do have major effects but that they are spread across so many different genes that the methodology applied here would fail to detect them.

### Limitations

It is possible that at least some participants who reported seeing a psychiatrist had in fact instead seen a psychologist or counsellor. The phenotype does not clearly correspond to a single psychiatric diagnosis but is intended to pick up some broad level of psychiatric morbidity, such as is met by slightly more than 10% of the sample. No attempt was made to exclude psychiatric disorder in controls and it is likely that some will have had significant morbidity.

## Conclusion

As for the smaller sample, this study completely fails to identify genes which might influence the risk of mental disorder of sufficient severity to warrant referral for psychiatric treatment. Given the complete lack of signal, there is no expectation that simply increasing the sample size would yield useful results. Instead, progress in understanding the genetics of affective disorders might depend on obtaining sequence data from large samples of more severe and narrowly defined phenotypes, such as bipolar 1 disorder or severe depression requiring repeated hospitalisation.

## Data Availability

The raw data is available on application to UK Biobank. Detailed results with variant counts cannot be made available because they might be used for subject identification.

## Acknowledgments

This research has been conducted using the UK Biobank Resource. The author wishes to acknowledge the staff supporting the High Performance Computing Cluster, Computer Science Department, University College London. This work was carried out in part using resources provided by BBSRC equipment grant BB/R01356X/1.

